# Mystical Experience Induced by Esketamine Treatment: A Real-World Observational Study

**DOI:** 10.64898/2026.03.31.26349757

**Authors:** Maia Mallevays, Louise Fuet, Michel Danon, Laura Di Lodovico, Claire Jaffré, Louise Bouzeghoub, Seifeddine Mrad, Anne-Victoire Rousselet, Lola Allary, Chlöé Müh, Bente Vissel, Pierre De Maricourt, Fabien Vinckier, Raphaël Gaillard, Lila Mekaoui, Philip Gorwood, Anne-Cécile Petit, Lucie Berkovitch

**Affiliations:** Cognitive Neuroimaging Unit, INSERM, CEA, Université Paris-Saclay, NeuroSpin center, 91191 Gif/Yvette, France; Institut de Neuromodulation, GHU Paris Psychiatrie et Neurosciences, Hôpital Sainte-Anne, pôle hospitalo-universitaire 15, Université Paris Cité, Paris, France; Université Paris Saclay, École Doctorale BioSigN (BioSigne), Life Sciences and Health Graduate School, 91190/91191 Gif-sur-Yvette, France; Service Hospitalo-Universitaire, Pôle Hospitalo-Universitaire Psychiatrie Paris 15, Hôpital Sainte Anne, GHU Paris Psychiatrie et Neurosciences, Paris; Clinique des Maladies Mentales et de l’Encéphale (CMME), Hôpital Sainte-Anne, GHU Paris Psychiatrie et Neurosciences, Paris, France; Université Paris Cité, F-75006 Paris, France; INSERM U1266. Institute of Psychiatry and Neuroscience of Paris (IPNP), Paris, France; Secteur 15-16, Pôle Hospitalo-Universitaire Psychiatrie Paris 15, Hôpital Sainte Anne, GHU Paris Psychiatrie et Neurosciences, Paris, France; Motivation, Brain & Behavior (MBB) lab, Paris Brain Institute, F-75013, Paris, France; Institut Pasteur, CNRS UMR 3571, Perception and Action Unit, F-75015 Paris, France; PsyBrain Team, UNIACT, NeuroSpin, CEA Saclay, Paris, France

**Author notes:** Corresponding author: Maia Mallevays, Institut de Neuromodulation, GHU Paris Psychiatrie et Neurosciences, Hôpital Sainte-Anne, 12 Rue Cabanis, Paris 75014, France. these authors equally contributed to the work (Maia Mallevays, Louise Fuet).

**Keywords:** Mystical experience, ketamine, esketamine, treatment-resistant depression, antidepressant

## Abstract

Esketamine is a fast-acting antidepressant drug which induces acute psychoactive effects. The most frequent is a dissociative state which seems unrelated to therapeutic efficacy. Other esketamine-induced effects, including psychedelic-like mystical experiences, have been poorly studied in terms of phenomenology and frequency, and may carry specific therapeutic relevance. In this study, we characterised esketamine-induced mystical experiences in relation with clinical outcomes. We conducted a longitudinal observational study and systematically measured acute subjective effects in patients receiving esketamine for treatment-resistant depression after each administration across the induction phase. A total of 45 patients were included, from two independent centres, totalling 352 esketamine administrations. Principal Component Analysis (PCA) supported the validity of the Mystical Experience Questionnaire (MEQ-30) for assessing esketamine-induced subjective effects, with components recovering dimensions previously validated with classic psychedelics. Mystical experiences (MEQ-30 score ≥ 60) occurred in 58% of patients, with high inter- and intra-individual variability in frequency, intensity, and phenomenology across sessions. Higher mean and peak MEQ scores were associated with greater improvement in Montgomery-Åsberg Depression Rating Scale scores from pre- to post-treatment, whereas the intensity of dissociative or other non-mystical effects was not. Positive mood and mystical MEQ dimensions in particular predicted therapeutic outcomes. Baseline spirituality also significantly predicted treatment outcomes and peak MEQ scores in the first week of treatment. These findings add to the growing body of evidence suggesting that psychedelic-like mystical experiences may be associated to therapeutic efficacy, not only in classic psychedelic-assisted therapy, but also in esketamine treatment.

## 1. Introduction

Treatment-resistant depression (TRD), defined as the failure to respond to at least two adequate treatment trials (1,2), affects at least 30% of patients with major depressive disorder (3,4) and approximately one-quarter of patients with bipolar disorder (5), posing a major public health challenge (6). Among emerging treatment strategies (7–9), ketamine and its enantiomer esketamine have demonstrated fast-acting antidepressant effects in TRD (10–25) and acute suicidal crises (26–34), leading to regulatory approval of intranasal esketamine for TRD in 2019 (35,36) and growing clinical use (37,38).

The primary subjective effect associated with (es)ketamine is dissociation, which encompasses derealisation, defined as a sense of unreality or detachment from one’s surroundings, and depersonalisation, defined as a disconnection from one’s own body, thoughts, or sense of self. However, its relationship with antidepressant efficacy has been inconsistent (39–43), suggesting dissociation is not a reliable predictor of antidepressant effects.

Psychedelic substances such as psilocybin and LSD have recently demonstrated promising fast-acting antidepressant effects (44,45), accompanied by strong acute subjective effects, including “mystical experiences” (46,47). Mystical experiences are characterised by a profound sense of interconnectedness with others or a larger whole (unity); a deep intuitive sense of understanding about oneself or reality (insight); a spiritually meaningful experience, often involving encounters with sacred or spiritual entities (spirituality); intense positive emotions (bliss); altered perceptions of time and space (transcendence); and a quality of ineffability that make these experiences difficult to explain with words (48). In clinical trials using psychedelic-assisted therapy, mystical experience intensity has emerged as a predictor of therapeutic response across multiple conditions, including TRD, addictive disorders and depressive and anxiety symptoms in patients with life-threatening diseases (49–51). These associations have been linked specifically to dimensions of “oceanic boundlessness”, encompassing bliss, unity and insight (52), and transcendence of time and space (53), as measured by established scales such as the Five-Dimensional Altered States of Consciousness (5D-ASC), 11-Dimensional Altered States of Consciousness (11D-ASC) and the 30-item Mystical Experiences Questionnaire (MEQ-30) (54–57).

Though far less studied, emerging evidence suggests that (es)ketamine also induces mystical experiences that may contribute to its therapeutic efficacy (41,58–60). A placebo-controlled randomised trial observed that ketamine’s antidepressant effects correlated with 11D-ASC dimensions of spirituality, unity and insight (58). More recently, ketamine’s antidepressant effects have been linked to MEQ-measured mystical experience intensity (59) and ego dissolution (60). Notably, Aepfelbacher et al. (41) found that scores on the Awe Experience Scale (AWE-S), which captures an emotional state wherein one’s usual frame of reference is challenged by something vast or profound, mediated the relationship between ketamine administration and reductions in depressive symptoms in a placebo-controlled randomised trial for TRD. However, the relationship between mystical experiences and clinical outcomes has not been consistently replicated. Aust et al. (61) found no difference in 5D-ASC oceanic boundlessness scores between treatment responders and non-responders, with response defined as a ≥50% depression score reduction. Notably, in that study, mystical effects and treatment response were assessed only once, 24 hours after the first ketamine infusion, preventing to capture potential antidepressant effects across subsequent sessions.

In sum, while preliminary evidence links mystical experiences to (es)ketamine’s therapeutic efficacy, empirical data remain limited and inconsistent. Importantly, very few studies have systematically investigated (es)ketamine-induced mystical experiences in ecological and real-life conditions. As MEQ-30 and 5/11D-ASC scales were developed for classic psychedelics, their suitability for capturing (es)ketamine-induced subjective effects remains uncertain.

The present study has three aims: (1) to assess whether psychedelic experience questionnaires accurately capture esketamine-induced acute subjective effects; (2) to document the occurrence, phenomenology, and frequency of esketamine-induced mystical experiences; (3) to explore the relationship between esketamine’s acute subjective effects and antidepressant outcomes. To this end, we prospectively measured acute subjective effects in all eligible patients receiving esketamine for TRD in routine clinical care, after each esketamine administration across the first month of treatment.

## 2. Methods

### 2.1. Study design

This observational study collected data from a naturalistic cohort of patients in two different units at the GHU Paris Psychiatrie et Neurosciences (*Clinique des Maladies Mentales et de l’Encéphale* and the *Service Hospitalo-Universitaire* at Sainte-Anne Hospital in Paris) between June 2024 and April 2025.

The inclusion criteria were diagnosis of unipolar or bipolar TRD with an indication for esketamine. Exclusion criteria included a switch to intravenous administration within the induction phase. Participants were enrolled before the first treatment session. Classical time-course of esketamine treatment induction consists of 8 administrations on a twice-weekly basis. The follow-up period lasted four weeks, with the option to extend to five weeks if the induction phase was prolonged. Data collection occurred at each administration, i.e., twice weekly per participant.

### 2.2. Power analysis

A power analysis is provided in Supplementary Materials.

### 2.3. Regulatory

This study was conducted in accordance with the declaration of Helsinki and the Jardé law. It was registered on the Health Data Hub under the number 27444021. This research was approved by the GHU Paris Research Ethics Committee (accreditation number 2025-CER-A-016). All patients received detailed oral and written information regarding study objectives and procedures and provided written informed consent before enrolment.

### 2.4. Measures

#### 2.4.1. Sociodemographic and clinical characteristics

Sociodemographic data, medical history, resistance profile and indication for esketamine treatment were collected by the clinical team and supplemented through patient interviews. Treatment resistance was graded according to the Thase & Rush (1997) staging model for unipolar patients (62).

#### 2.4.2. Clinician-Administered and Self-Report Assessment of Depressive Symptoms and **Suicidality**

Depressive symptoms were assessed using the Clinical-Administered Montgomery-Åsberg Depression Rating Scale (MADRS) (63) and the Quick Inventory of Depressive Symptomatology-Self-Report (QIDS-SR16) (64). Suicidal ideation and behaviour were assessed at baseline using the Columbia-Suicide Severity Rating Scale (C-SSRS) (65). A baseline assessment (QIDS-SR16, MADRS, C-SSRS) was conducted prior to initiation of esketamine treatment. Subsequent MADRS assessments were performed prior to each treatment session, thereby reflecting the effect of the previous session. A final assessment (MADRS and QIDS-SR16) was completed within one week following the end of the induction phase (8 treatment sessions administered twice weekly, see Figure 1). Changes in depression symptoms post-treatment were operationalised as the difference between baseline MADRS and MADRS score after the final induction session (ΔMADRS = baseline - post-treatment).

**Figure 1.**
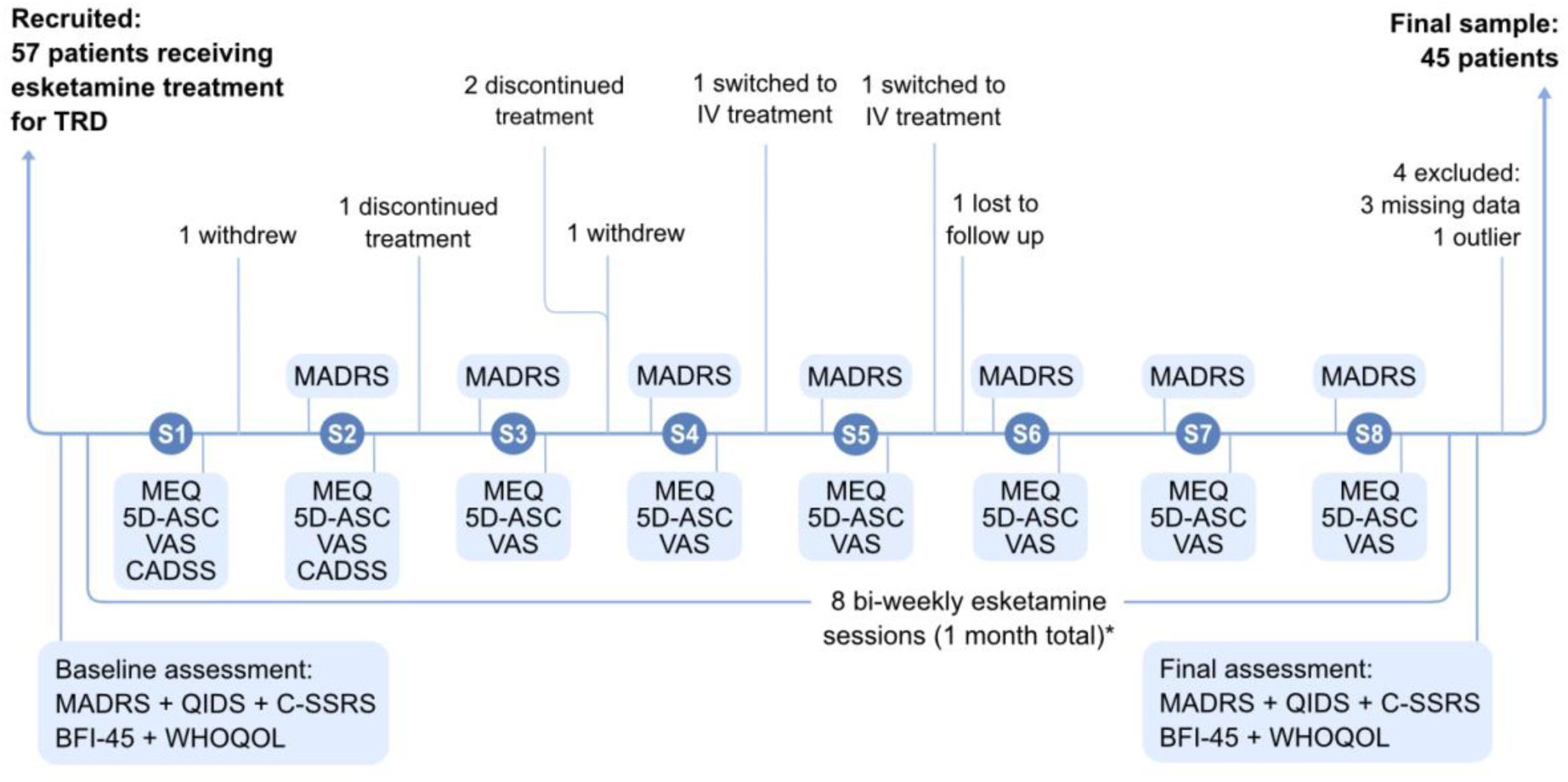
Flow of participants and assessment schedule. Baseline assessments were completed <48h before the first session, and final assessments within 1 week after the end of the 8 induction sessions, before the next esketamine session if applicable. Clinician-rated MADRS was administered before each session, thereby reflecting the effect of the previous session, and patients completed the MEQ (plus 5D-ASC when MEQ≥45), and the VAS (n = 29 completed the VAS scale) within 24h after each session (usually a couple of hours after the session). Dissociation score data (CADSS or 0-5 dissociation intensity) was collected for assessments done after session 1 and session 2. TRD = Treatment-Resistant Depression; S1-S8 = Sessions 1-8 of esketamine treatment; MADRS = Montgomery-Åsberg Depression Rating Scale; QIDS = Quick Inventory of Depressive Symptomatology; C-SSRS = Columbia-Suicide Severity Rating Scale; BFI-45 = Big Five Inventory-45; WHOQOL = World Health Organization Quality of Life questionnaire; MEQ = Mystical Experience Questionnaire; 5D-ASC = 5-Dimensional Altered States of Consciousness Rating Scale; IV = Intravenous ketamine.

#### 2.4.3. Subjective Effects During Treatment

Mystical experiences were assessed with the Mystical Experience Questionnaire (MEQ-30), completed by patients within 24 hours after each treatment session (Figure 1). The MEQ-30 is organised into four dimensions: Mystical, Positive Mood, Transcendence of Time and Space, and Ineffability (54,56,66). We adopted a threshold value of 60 points (40% of the maximum score) to delineate two subgroups: “with mystical experience” (MEQ-30 ≥ 60 at least once during treatment) and “without mystical experience.” (Supplementary for details).

To further characterise subjective experiences in sessions showing at least minimal mystical-type effects, we collected the Five-Dimensional Altered States of Consciousness (5D-ASC) in sessions where MEQ-30 scores exceeded 45. This questionnaire comprises 94 items across five dimensions (Oceanic Boundlessness, Anxious Ego Dissolution, Visual Alterations, Auditory Alterations, Vigilance Reduction), each on a Likert scale from 0-100 (55).

To investigate the emotional valence and intensity of non-mystical subjective experiences, Visual Analogue Scales (VAS) similar to that used in previous studies investigating drug-induced subjective experiences (67–70) were added to the protocol after recruitment began and completed within 24 hours after each session.

Clinician-rated dissociation scores were collected retrospectively from the medical database. All available dissociation ratings in sessions 1 and 2, including the Clinician-Administered Dissociative States Scale (CADSS) and clinician-rated dissociation intensity (0-5 intensity score), were included in the analysis (Supplementary Methods).

#### 2.4.4. Spiritual wellbeing and Big Five personality

Spiritual wellbeing was assessed using the World Health Organization Quality of Life Spirituality, Religion and Personal Beliefs questionnaire (WHOQOL-SRPB), selected for its validation in French and applicability beyond religiosity (71,72). All participants completed this questionnaire at baseline and within the week following the end of the induction phase. Personality traits were measured using the French version of the Big-Five Inventory-45 (BFI-45) (73) with five dimensions (Openness, Conscientiousness, Extraversion, Agreeableness, and Neuroticism) administered at baseline and at the end of the induction phase (Figure 1).

### 2.5. Principal Component Analyses

Since the scales used to measure subjective effects are not validated for esketamine, we applied Principal Component Analyses (PCA) to identify data-driven dimensions of esketamine’s subjective effects. PCA were run separately on all MEQ-30 items and all VAS items using a methodology similar to previous studies (74–78) (Supplementary Methods). A PCA combining all MEQ-30 and VAS items can be found in Supplementary Materials (Figure S4).

### 2.6. Statistical Analysis

All statistical analyses were performed using R statistical software (https://www.r-project.org). To correct for multiple comparisons, we applied Bonferroni corrections or the Benjamini-Hochberg (BH) procedure where applicable. To explore the absence of differences, we ran Bayesian statistics using the BayesFactor package (79).

Analyses of variance (ANOVA) controlling for age, sex, diagnosis (unipolar vs. bipolar), and treatment centre (SHU vs. CMME) were conducted to test whether (a) measures of subjective experience predicted antidepressant response and (b) baseline measures of WHOQOL spirituality and Big Five personality predicted antidepressant response or mystical experiences.

Pearson correlations were used between subjective experience scores. T-tests were applied to measure changes in WHOQOL spirituality and Big Five personality scores from pre- to post-treatment (Supplementary Methods).

## 3. Results

### 3.1. Participants

Fifty-seven participants were recruited, of whom 12 were excluded (Figure 1; Supplementary Methods), yielding a final sample of 45 participants.

### 3.2. Socio-demographic characteristics

Socio-demographic characteristics are described in Table 1.

**Table 1.** Sociodemographic and clinical characteristics.

| Characteristic | N = 45<sup>1</sup> |
| --- | --- |
| **Age (years)** | 46.36 (15.00) |
| **Centre** |  |
| *CMME* | 23 (51%) |
| *SHU* | 22 (49%) |
| **Gender** |  |
| *F* | 25 (56%) |
| *M* | 20 (44%) |
| **Education level** |  |
| *Less than high school* | 3 (6.7%) |
| *High school diploma* | 4 (8.9%) |
| *Bachelor's degree* | 8 (18%) |
| *Bachelor's to master's degree* | 21 (47%) |
| *>Postgraduate degree (Master's/Doctorate)* | 9 (20%) |
| **Socio-professional category** |  |
| *Employees/Clerical/Office Workers* | 6 (13%) |
| *Manual Laborers* | 1 (2.2%) |
| *Retirees* | 5 (11%) |
| *Senior Officials/Managers/Executives* | 16 (36%) |
| *Skilled Manual Workers* | 2 (4.4%) |
| *Students* | 5 (11%) |
| *Technicians/Associate Professionals* | 7 (16%) |
| *Unemployed/Not in Labor Force/Economically Inactive* | 3 (6.7%) |
| **Mood disorder type** |  |
| *Bipolar* | 11 (24%) |
| *Unipolar* | 34 (76%) |
| **Baseline MADRS total score** | 27.27 (5.78) |
| **Baseline QIDS total score** | 18.30 (4.42) (missing 1) |
| **Baseline C-SSRS suicide score** | 1.02 (1.14) |
| **Baseline WHOQOL spirituality** | 61.40 (24.59) |
| **Episode duration (months)** | 38.00 (47.46) (missing 4) |
| **Resistance level (Thase & Rush staging for unipolar depression)** | 2.65 (0.95) (missing 12) |
| **Number of past hospitalisations** | 2.42 (1.97) (missing 1) |
| **Number of past suicidal attempts** | 0.61 (1.15) |
| **Lifetime addictive disorder** |  |
| *No* | 29 (64%) |
| *Yes* | 16 (36%) |
| **Personality disorder diagnosis** |  |
| *No* | 40 (89%) |
| *Yes* | 5 (11%) |

<sup>1</sup>Mean (SD); n (%); (missing data for n participants)

### 3.3. Esketamine treatment course

Most patients received 8 biweekly sessions during the induction phase (two received 9 sessions and 3 were assessed after 7; Supplementary Methods) totalling 352 administrations. Regarding initial dose, 19 participants started at 84 mg, 21 at 56 mg, and 5 at 28 mg (Supplementary Methods).

#### 3.3.1. Antidepressant effects of esketamine

First, we confirmed esketamine’s antidepressant effects in our sample: 49% of patients (n = 22) responded to treatment (i.e., had ≥ 50% MADRS reduction from baseline to post-treatment), and 38% (n = 17) showed remission (i.e., had a final MADRS score ≤ 10). MADRS and QIDS scores were significantly reduced at the end of the induction phase (MADRS mean decrease: 12.07 point; *t*(44) = 10.10, *p* < 0.001; QIDS mean decrease: 4.55 points; *t*(41) = 5.43, *p* < 0.001).

Antidepressant effects appeared within the first week of treatment (after two sessions; mean MADRS change = −6.45 ± 6.16, *t*(43) = −6.95, *p* < 0.001). Improvement at one week predicted overall response (F(5,38) = 2.77, *p* = 0.03).

### 3.5. Mystical effects of esketamine

We then characterised acute subjective effects during esketamine administrations. First, we measured mystical experience using the MEQ after each session. Across participants and sessions, the mean MEQ score was 36.6/150 (SD = 27.7), with a mean peak score of 61.6/150 (SD = 32.4), defined as the maximum score per participant. Using a threshold of MEQ ≥ 60, 22% of sessions met criteria for a mystical experience, and 26 participants (58%) experienced at least one mystical session. Within this mystical group, such experiences occurred in 38% of sessions, with a mean score of 51.7/150 (SD = 26.6) and a mean peak of 85.1/150 (SD = 20.2). In contrast, the non-mystical group had a mean score of 16.2/150 (SD = 11.1) and a mean peak score of 29.5/150 (SD = 10.5). Figure 3a shows the trajectory of mean MEQ scores across sessions for each group.

The intensity of mystical experiences was highly variable across sessions (Figure S1).

PCA identified 3 significant data-driven components of esketamine-induced mystical experience. The first principal component (PC) primarily reflected the mystical dimension, paired with partial loadings on transcendence of time and space, as well as ecstasy and awe. The second PC reflected co-occurring ineffability and transcendence of time and space. The third PC reflected positive mood (Figure 2 c-f). As these PCs closely map onto the previously validated MEQ dimensions, our findings support the use of this scale for characterising esketamine-induced subjective effects, and pre-defined MEQ dimensions are used in subsequent analyses.

**Figure 2.**
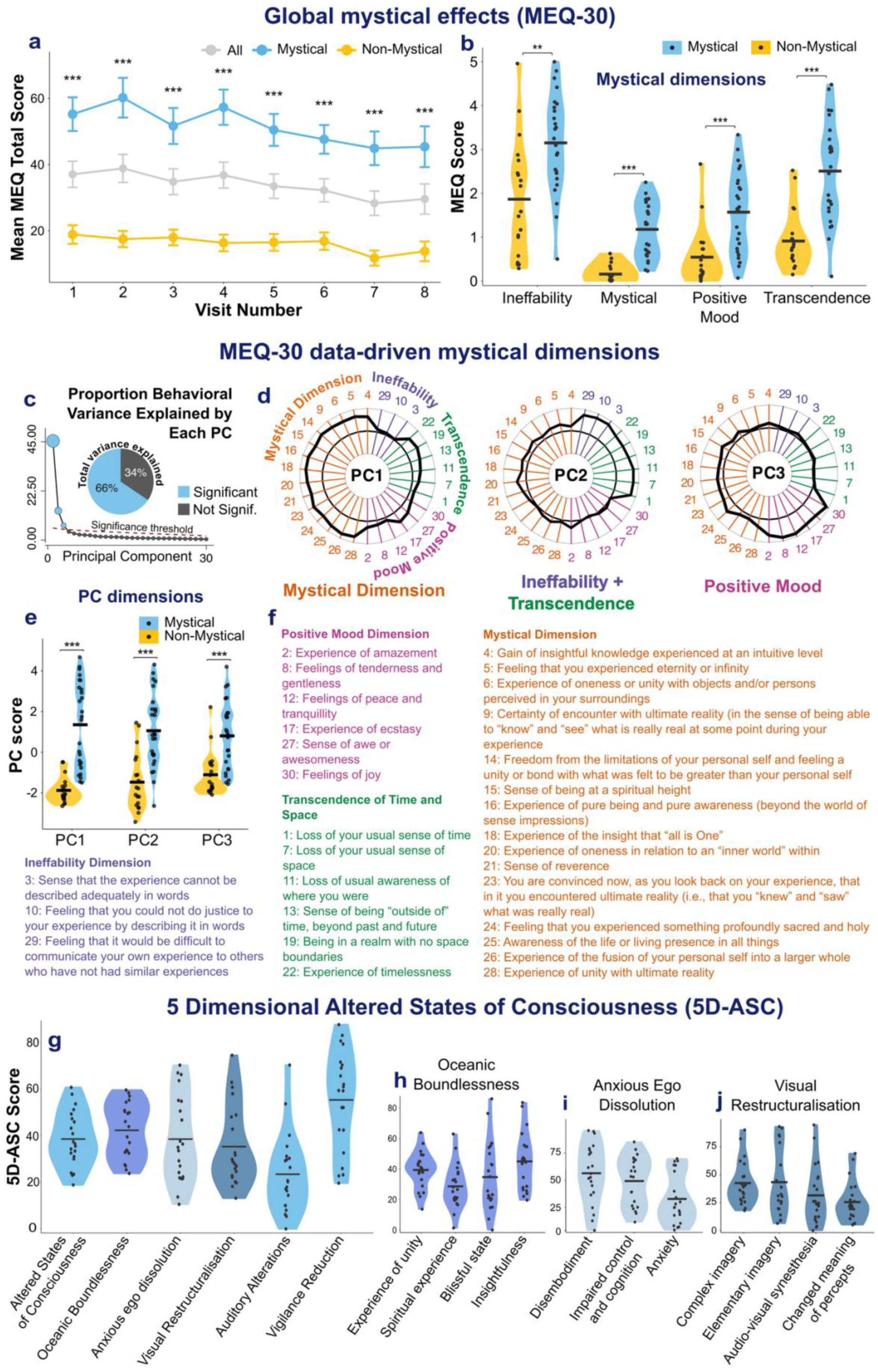
Characterising esketamine-induced mystical effects. (a) Mean MEQ-30 total scores (0–150) across treatment sessions for the whole population (grey), mystical group (blue), and non-mystical group (yellow). The mystical group was defined as participants who scored ≥ 60 on the MEQ-30 in at least one session. (b) Violin plots showing MEQ dimension scores (Ineffability, Mystical, Positive Mood, Transcendence of Time and Space) in mystical (blue) and non-mystical (yellow) groups. Mean scores were significantly higher in the mystical group across all dimensions. (c) Scree plot showing the percentage of variance explained by each principal component (PC); PC1 explained 45%, PC2 explained 13%, and PC3 explained 7%, with significant PCs together accounting for 66% of the total variance. (d) Radar plots showing PC loadings for each MEQ-30 item, corresponding to the data-driven mystical dimensions. (e) Violin plots showing PC scores by group (mystical vs non-mystical). (f) MEQ-30 items grouped by their four dimensions: Mystical, Positive Mood, Ineffability, and Transcendence of Time and Space. (g) Violin plots showing total scores and the five core dimensions of the 5D-ASC: Oceanic Boundlessness (OB), Anxious Ego Dissolution (AED), Visual Restructuralisation (VR), Auditory Alterations (AA), and Vigilance Reduction (VR), in participants who completed the 5D-ASC (n = 21). (h–j) Distributions of scores across the subdimensions of (h) OB (experience of unity, spiritual experience, blissful state, insightfulness), (i) VR (complex imagery, elementary imagery, audiovisual synaesthesia, changed meaning of percepts), and (j) AED (disembodiment, impaired control and cognition, anxiety). The highest 5D-ASC score was vigilance reduction, reflecting decreased alertness, mental slowing and drowsiness. Within oceanic boundlessness, the most strongly expressed subdimensions were experience of unity and insightfulness. Anxious ego dissolution was primarily driven by disembodiment, indicating a sense of detachment from the physical body, while anxiety-related items were less prominent overall. Visual restructuralisation was present to a lesser extent and mainly reflected changes in imagery rather than complex visual hallucinations.

Mean MEQ dimension scores were greater in the mystical compared to the non-mystical group for all dimensions (all Bonferroni corrected *ps* < 0.001; Figure 2b).

In sessions with MEQ-30 scores ≥ 45, the 5D-ASC was also collected to obtain a more in-depth description of acute subjective effects. This occurred in 21 participants, all of whom belonged to the mystical group (defined by at least one session with MEQ-30 ≥ 60). The total mean 5D-ASC score was 38.45/100 (SD = 11.66), and the mean peak score was 45.68/100 (SD = 14.45). See Figure 2g-j for the distribution of 5D-ASC dimensions.

### 3.6. Visual Analogue Scales (VAS) and dissociation measures

Twenty-nine participants completed the VAS after each session, measuring affective valence and cognitive functions (Figure 3a and 3b). VAS item means did not show any significant differences between mystical and non-mystical groups (Supplementary Figure S2).

**Figure 3.**
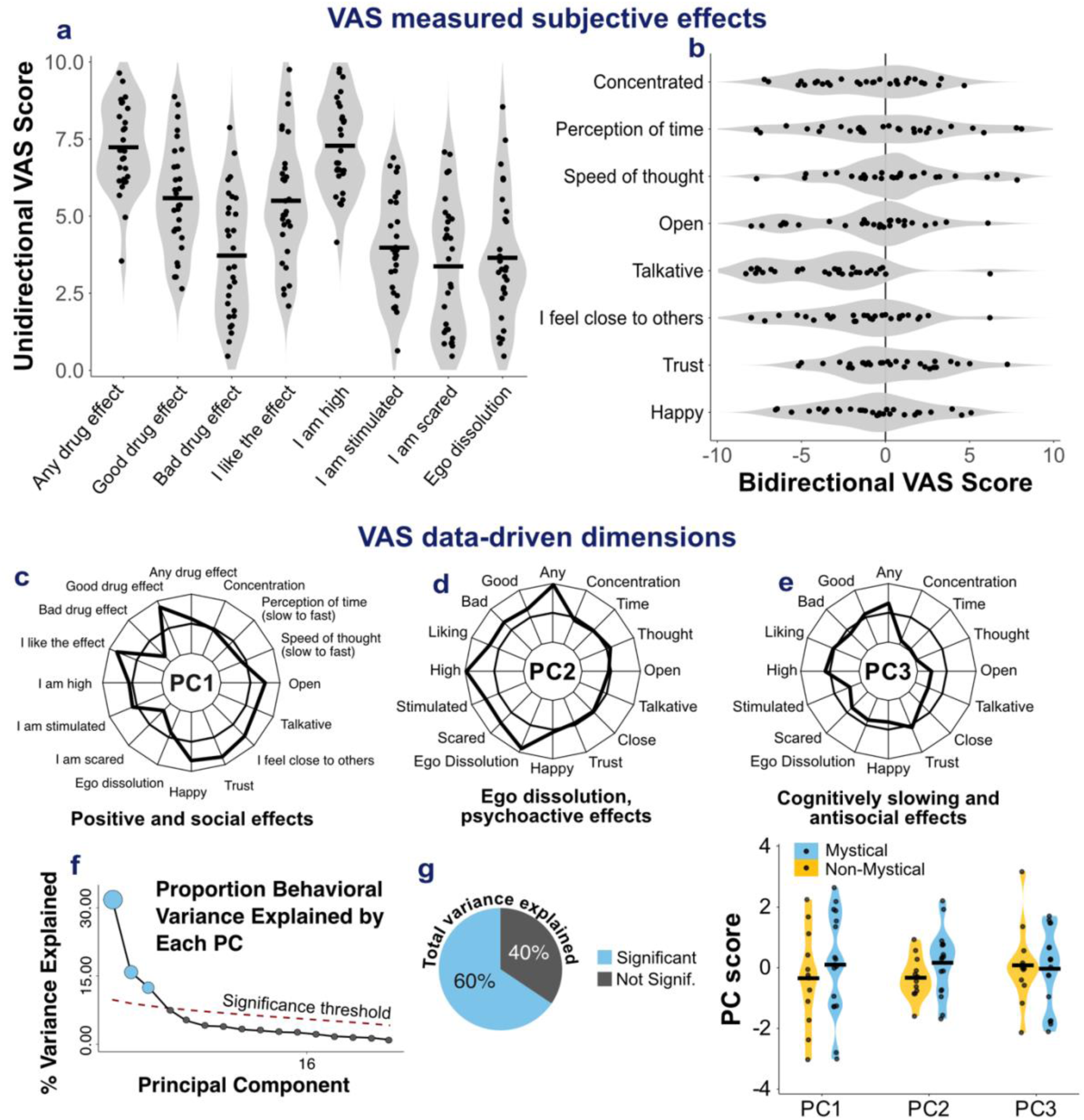
Characterising esketamine-induced valence-related and cognitive subjective effects. (a-b) Violin plots showing distributions of (a) unidirectional and (b) bidirectional VAS ratings in participants who completed the VAS scale (n = 29). The items “I am high”, “any drug effect”, “good drug effect”, “I like the effect”, showed the highest mean scores. (c-e) Radar plots showing principal component (PC) loadings reflecting data-driven dimensions of VAS responses: (c) PC1, positive and social effects; (d) PC2, ego dissolution, stimulant and scary effects; and (e) PC3, cognitively slowing and antisocial effects. (f-g) Scree plots showing the proportion of behavioural variance explained by each PC (f) and cumulative variance explained by significant components (g). PC1, PC2, and PC3 together explained 60% of the total variance (32%, 16%, and 12%, respectively). (h) Violin plots showing PC scores by group, comparing participants with mystical (blue) and non-mystical (yellow) experiences.

PCA analysis with all VAS item scores across sessions identified three dimensions of VAS-measured esketamine phenomenology. PC1 reflected positive emotional and social effects. PC2 captured ego dissolution effects, along with stimulation and fear. PC3 reflected cognitive slowing, paired with decreased sociality (Figure 3c-h).

First week dissociation scores were collected in 30 participants (mean = 3.32/5, SD = 1.58).

### 3.7. Relationships between acute subjective effects

We first studied the relationships between MEQ dimensions and VAS PC. Positive mood, as captured by both the positive mood MEQ dimension and VAS PC1, was correlated across the two measures (*r* = 0.66, BH corrected *p* < 0.001). Ego dissolution, captured by VAS PC2, correlated with the ineffability MEQ dimension (*r* = 0.67, BH corrected *p* < 0.001), and the transcendence of time and space MEQ dimension (r = 0.68, BH corrected *p* < 0.001). None of the other MEQ dimensions correlated with VAS PCs (all 1/BF > 1.05).

To explore the relationship between dissociation intensity and other measures, we restricted subsequent analyses to first-week scores, as dissociation was only collected during this period. First-week dissociation intensity was correlated with the feeling of transcendence of time and space, anticorrelated with positive mood and associated with cognitive slowing. Indeed, mean dissociation scores correlated with MEQ transcendence scores (*r* = 0.55, Bonferroni corrected *p* = 0.007). Mean dissociation scores did not significantly correlate with other MEQ dimensions, nor with mean MEQ scores (all *ps* > 0.14; all 1/BF > 0.68). Mean dissociation scores negatively correlated with VAS PC1 scores, reflecting positive and social effects (*r* = −0.69, Bonferroni corrected *p* = 0.006). Further, mean dissociation correlated with VAS PC3 scores, reflecting cognitive slowing and reduced sociality, but this association did not survive Bonferroni correction (r = −0.47, uncorrected *p* = 0.02, Bonferroni corrected *p* = 0.06). No significant correlation was observed between dissociation and VAS PC2 scores, reflecting ego dissolution (1/BF = 1.31).

### 3.8. Mystical experiences predict antidepressant effects of esketamine

We tested whether esketamine-induced acute subjective effects predicted clinical improvement.

Importantly, mystical experience intensity across sessions significantly predicted MADRS decrease from pre- to post-treatment after controlling for age, sex, diagnosis (unipolar vs. bipolar), and centre (none of these covariates had significant main effects). Higher peak MEQ scores predicted greater symptom reduction (F(1,39) = 5.91, *p* = 0.02), and this result was confirmed with mean MEQ scores (F(1,39) = 5.17, *p* = 0.029) (Figure 4a and 5c). Among participants who had at least one mystical experience, the frequency of mystical experiences was not associated with ΔMADRS (F(1,20) = 3.27, *p* = 0.09), suggesting that symptom improvement is not related to an accumulation of mystical experiences.

**Figure 4.**
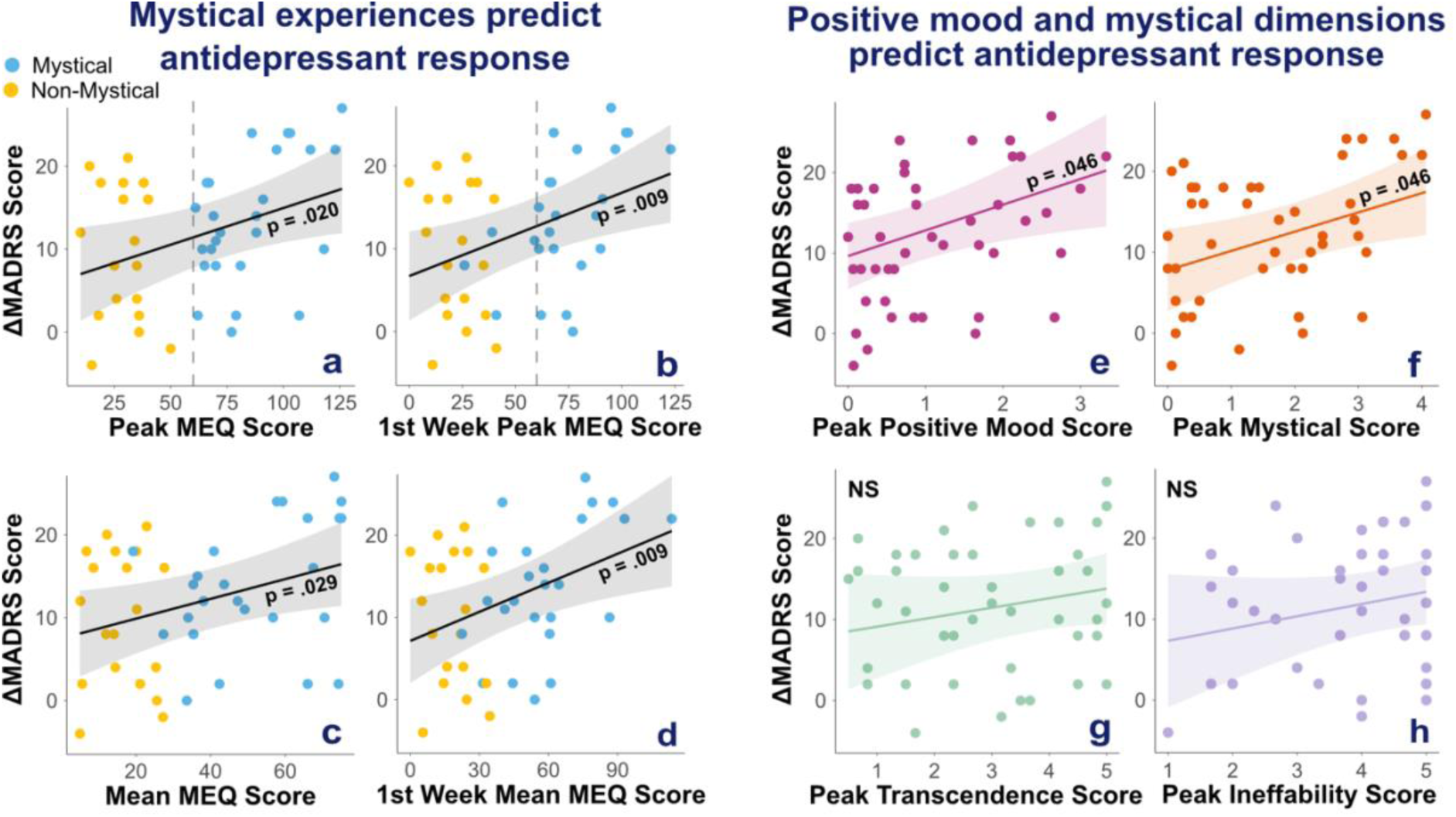
Mystical experience intensity predicts stronger antidepressant effects of esketamine. Regression lines represent predicted values from adjusted linear models in which MEQ-derived measures predict antidepressant response (ΔMADRS = pre-treatment minus post-treatment), controlling for age, sex, diagnosis (unipolar vs bipolar), and treatment centre (SHU vs CMME). Shaded areas represent 95% confidence intervals. Panels (a–d): participants are coloured by group (blue: mystical; yellow: non-mystical). Predictors were: (a) peak MEQ-30 scores across all sessions, (b) peak MEQ-30 scores during the first week (sessions 1–2), (c) mean MEQ-30 scores across all sessions, and (d) mean MEQ-30 scores during the first week. Panels (e–h): positive mood (e) and mystical dimension scores (f) predicted stronger antidepressant effects, whereas transcendence of time and space (g) and ineffability (h) did not. P-values were corrected using the Benjamini–Hochberg false discovery rate procedure across peak and mean subdimension scores (8 tests).

**Figure 5.**
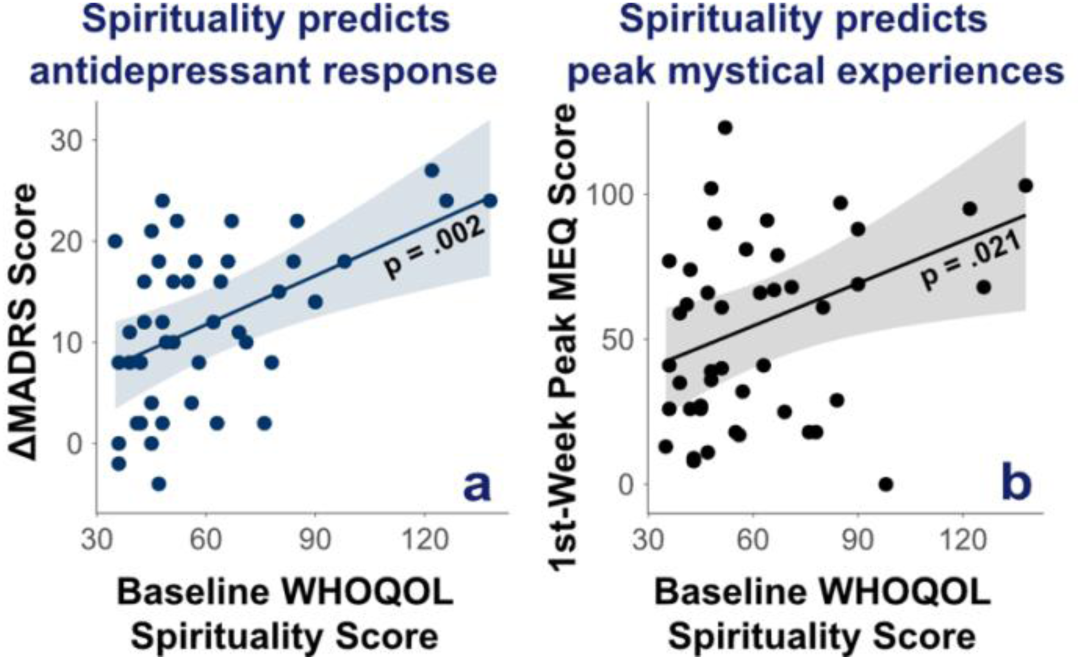
Baseline WHOQOL Spirituality scores predict greater antidepressant effects of esketamine and greater intensity of peak mystical experiences in the first week of treatment. Regression lines represent predicted values from adjusted linear models in which baseline total WHOQOL Spirituality scores predict (a) antidepressant response (ΔMADRS = pre-treatment minus post-treatment) or (b) first week peak MEQ scores, controlling for age, sex, diagnosis (unipolar vs bipolar), and treatment centre (SHU vs CMME). Shaded areas represent 95% confidence intervals.

Moreover, mystical experience intensity was an early predictor of clinical response as mean and peak MEQ scores in the first week of treatment (1^st^ 2 sessions) significantly predicted ΔMADRS at the end of induction (peak: F(1,39) = 7.58, *p* = 0.009; mean: F(1,39) = 7.66, *p* = 0.009) (Figure 5b and 5d).

By contrast, there was no immediate effect of mystical experience on depressive symptoms. Indeed, MEQ scores in the first week did not predict early improvement measured by ΔMADRS from baseline to post session 2 (peak: F(1,38) = 1.81, *p* = 0.19, 1/BF = 1.44; mean: F(1,38) = 2.71, *p* = 0.12, 1/BF = 1.08).

Similar results were observed with the self-report measures of depression (QIDS, see Supplementary Materials).

We then explored whether any specific dimension was associated with clinical improvement. We found that positive mood peak and mean scores predicted ΔMADRS (peak: F(1,39) = 6.20, BH corrected *p* = 0.046; mean: F(1,39) = 6.70, BH corrected *p* = 0.046). Mystical dimension peak scores also predicted ΔMADRS (F(1,39) = 6.27, BH corrected *p* = 0.046) whereas mean mystical scores did not (F(1,39) = 3.69, BH corrected *p* = 0.11, BF = 8.33). None of the other MEQ dimension peak or mean scores significantly predicted ΔMADRS (all BH corrected *ps* > 0.24, all 1/BF > 1.1).

In the 21 participants who completed the 5D-ASC scale, neither peak nor mean scores predicted ΔMADRS (peak: F(1,15) = 1.26, *p* = 0.28, 1/BF = 1.58; mean: F(1,15) = 0.82, *p* = 0.38, 1/BF = 1.6). Similarly, 5D-ASC dimensions were not significant predictors of ΔMADRS (all *ps* > 0.37).

### 3.9. Other acute subjective effects do not predict clinical improvement

Neither VAS nor dissociative measures significantly predicted ΔMADRS (VAS PC: all Bonferroni corrected *ps* > 0.63, all 1/BF > 0.57; dissociative scores peak and mean: all *ps* > 0.4, all 1/BF > 1.4).

### 3.10. Spirituality (WHOQOL)

Spirituality was assessed at baseline and post-induction to evaluate its evolution and its relationships with clinical improvement and acute subjective effects.

Spirituality did not change from pre- to post-treatment (pre-treatment mean WHOQOL: 61.40/160, SD = 24.59; post-treatment mean WHOQOL: 62.56/160, SD = 26.12; difference: *p* = 0.96). Baseline WHOQOL and baseline MADRS scores were not significantly correlated (*r* = 0.007, *p* = 0.96, 1/BF = 2.99).

Baseline WHOQOL scores significantly predicted both MADRS decrease across sessions (F(1,39) = 11.34, *p* = 0.002) and peak MEQ scores in the 1^st^ week of treatment (F(1,39) = 5.75, *p* = 0.021), but not other MEQ measures (all *ps* > 0.08 and 1/BF > 0.4) (Figure 5).

Baseline WHOQOL scores did not significantly predict VAS PCs (all *ps* > 0.25, all 1/BF > 1.19), or first week dissociation mean or peak (all *ps* > 0.28, all 1/BF > 1.36).

### 3.11. Big Five personality (BFI-45)

Finally, we explored how personality interacted with esketamine-induced antidepressant response and evolved with treatment.

First, we evaluated whether BFI-45 personality scores at baseline predicted ΔMADRS, mystical experience metrics, dissociation intensity or VAS PC scores, and found no significant results (all *ps* > 0.12, all 1/BF > 1.45).

Then, we studied personality changes after esketamine treatment and observed a decrease in Agreeableness (*ΔM* = −0.18, Bonferroni corrected *p* = 0.03), and in Neuroticism (*ΔM* = −0.25, Bonferroni corrected *p* = 0.02) (see Supplementary Results).

## 4. Discussion

### 4.1. Characterisation of esketamine-induced acute subjective effects

This study examined esketamine’s acute subjective effects and their relationship with treatment outcomes in patients with TRD. Esketamine produced a significant antidepressant response, with half of patients classified as responders after the induction phase. Mystical experiences (MEQ scores ≥ 60) occurred in over half of patients. PCA recovered previously validated MEQ dimensions, with PC1 reflecting the mystical dimension, PC2 a combination of transcendence and ineffability, and PC3 positive mood. These findings suggest that esketamine induces phenomenological dimensions of mystical experience which can be assessed with the same scales as those used to describe psychedelic experiences. Similarly, 5D-ASC allowed to measure esketamine-induced altered states of consciousness with scores encompassing oceanic boundlessness, self-related alterations, and perceptual changes. Mean VAS scores indicated strong psychoactive effects (“Any drug effects”, “I am high”) along with an overall positive valence (“Good drug effects”, “I like the effect”). Dissociation intensity was associated with reduced positive affect (VAS PC1), whereas mystical experiences were not, suggesting that dissociation may be experienced as aversive, while mystical experiences vary in emotional valence. Together, these findings show that mystical experiences are frequent during esketamine treatment and can be captured with scales used for psychedelic drugs.

### 4.2. Acute subjective effects and antidepressant efficacy

The role of the psychedelic experience in clinical outcomes remains debated (80,81). In serotonergic psychedelic therapy, mystical dimensions such as unity, spirituality, and bliss are associated with antidepressant outcomes (49,51,52,82), though not all studies replicate this finding (83,84). Preclinical work using antidotes showed that antidepressant effects can occur without psychedelic experience (85,86), and a case-report of TRD found that blocking psilocybin’s acute effects did not reduce efficacy (87).

As for ketamine, acute subjective effects are usually framed as side effects, using measures of dissociation or psychotomimetic effects (88). Our findings challenge this framing, showing that esketamine can also induce psychedelic-like mystical experiences, which may be clinically relevant and orthogonal to dissociative effects. Consistent with prior work (41,58,59), mystical experience intensity overall and during the first week predicted antidepressant effects across the full treatment course. In particular, MEQ positive mood and mystical dimensions predicted antidepressant effects, in line with prior studies linking treatment response with spirituality, unity, insight, ego dissolution and awe in both (es)ketamine treatment (41,58,60) and psychedelic-assisted therapy (52,89). Nevertheless, and consistent with previous results (61), mystical experience intensity did not predict early antidepressant response after one week, suggesting that mystical experiences may require time to exert an effect on depression.

By contrast, dissociation and non-mystical subjective effects did not significantly predict clinical outcomes in our study, in line with existing data (42,90,91). Similarly, 5D-ASC total scores did not predict treatment response, potentially due to ceiling effects as this scale was only collected in the mystical group.

Taken together, converging evidence supports a specific role for mystical or awe-related experiences, rather than dissociative effects or general psychoactive intensity, in ketamine’s antidepressant action. Mystical states may play a role in therapeutic outcomes by promoting psychological shifts and emotional breakthroughs. Qualitative reports support this, describing ketamine-induced mystical experiences that promote “letting go”, decentring from ruminative, self-referential thinking, and promoting connection with others, and with a great whole (58). Such experiences may open a window of cognitive flexibility and heightened sensitivity to positive emotions thereby favouring an update towards positive mental representations (92–94).

### 4.3. Spirituality, mystical experiences and antidepressant effects

Higher baseline spirituality predicted both the intensity of mystical experiences during the first week and greater clinical response. This is consistent with previous findings showing that individuals with stronger spiritual orientations were more likely to report mystical-type experiences during psychedelic states (95,96), and that baseline spirituality predicted better treatment response in depression (97,98). The form of spirituality assessed here, and in prior studies reporting similar findings, refers to “intrinsic” spirituality, which is centred on personal meaning, purpose, and a felt connection to something larger, rather than “extrinsic” religiosity, focused on outward religious practices (97,98). Intrinsic spirituality may serve as a coping resource and a source of resilience and meaning in depression (99–101).

### 4.4. Strengths and limitations

Several limitations should be noted. First, the observational, unblinded design introduces potential self-selection, expectancy, and suggestibility biases. Second, because data were collected in routine care within a naturalistic patient sample, assessments could not be fully standardised, despite efforts to keep them comparable across patients. In particular, CADSS was collected inconsistently and merged with a related but distinct dissociation scale, making dissociation findings exploratory at best. In addition, concomitant antidepressant treatments varied and were not controlled. Third, sample size was small for some measures (VAS: n = 29, 5D-ASC: n = 21), reducing power.

Nevertheless, this is one of the first studies to characterise acute subjective experiences during esketamine treatment, with particular attention to understudied mystical-type experiences. Combining complementary measures (MEQ, 5D-ASC, VAS) provided a broader perspective, and repeated assessments across sessions captured the frequency, intensity, and phenomenology of mystical experiences during the induction phase.

### 4.5. Perspectives

The role of the clinical setting remains a key open question. While several studies report that elements such as music or supportive environments enhance mystical experiences under (es)ketamine, their therapeutic relevance remains mixed (59,102,103). Relatedly, emerging studies are investigating combinations of (es)ketamine with psychotherapy, proposing that this could create a synergistic dynamic, with (es)ketamine opening patients to new perspectives and priming them for therapeutic engagement, while psychotherapy could support healing pathological representations and consolidate long-term change (60,104,105).

Investigating the neurobiological basis of ketamine’s acute subjective effects, particularly mystical experiences, would allow for a deeper understanding of their role in therapeutic outcomes. The insula and default mode network have been implicated in feelings of bodily harmony, ego-dissolution and spiritual insight (106–114). Positive affective components of mystical experiences have been linked to altered amygdala responses to emotional stimuli (115–117). Finally, neurocomputational hypotheses suggest that NMDA receptor blockade may transiently relax rigid priors within predictive processing frameworks, leading to more flexible mental states, and alter interoceptive processing, contributing to changes in bodily self-representation (93,118).

### 4.6. Conclusion

Our study is one of the first to investigate and characterise mystical-type experiences induced by esketamine in routine clinical care, following patients over the whole induction phase of treatment. Findings show that esketamine-induced mystical-type experiences are frequent in patients with TRD and can be measured with existing scales designed or used in psychedelic studies. Further, the intensity of such mystical experiences, as well as patients’ spirituality prior to treatment onset, were associated with esketamine’s antidepressant effects. These findings highlight the relevance of taking these experiences into account when administering (es)ketamine, for example by adapting the clinical setting. A deeper understanding of the neurobiological mechanisms and long-term psychological effects of these experiences could help rigorously characterise them, potentially broadening biomedical approaches to incorporate existential and spiritual dimensions of healing (119).

## Data Availability

The data supporting this study are not publicly available due to patient confidentiality.

## Acknowledgements

We would like to thank all patients who took the time to participate and complete the questionnaires.

Lucie Berkovitch thanks the Fondation Bettencourt-Schueller, the Philippe Foundation, the Foundation L’Oréal-Unesco and the National Institute of Mental Health (R01MH116038 and U01MH121766) for their support.

