## Supplementary Materials for "Mystical Experience Induced by Esketamine Treatment: A Real-World Observational Study"

### 6. Supplementary Materials

#### 6.1. Supplementary Methods

##### 6.1.1. Measures

*6.1.1.1. The Mystical Experience Questionnaire (MEQ-30)*

The MEQ-30 questionnaire has been validated across multiple psychometric studies, demonstrating strong reliability and validity in capturing the core characteristics of mystical experiences, particularly in the context of psychedelic research (1). It is widely used in clinical studies to assess the subjective effects of psychoactive substances and their therapeutic potential (2,3). The version used in this study was validated in French (4).

We adopted a threshold value of 60 points (40% of the maximum score) to delineate two subgroups: “with mystical experience” (MEQ-30 ≥ 60 at least once during treatment) and “without mystical experience.” Frequency was calculated as the percentage of sessions in which MEQ-30 score ≥ 60. This threshold was chosen based on previous classic psychedelic studies in which thresholds around 60% of the maximum score have been used to operationalise a “complete” mystical experience and was adjusted downward to account for the generally lower-intensity subjective effects induced by esketamine.

*6.1.1.2. The Five‑Dimensional Altered States of Consciousness (5D‑ASC) questionnaire*

The 5D-ASC has been widely used and validated in research exploring the effects of psychedelics on consciousness. It is regarded as a robust tool for capturing the complex and multifaceted aspects of psychedelic experiences. The scale has been adapted and applied across various research contexts to better understand subjective alterations in consciousness (5–9). A threshold of ≥ 60% on the “Oceanic Boundlessness” dimension was considered indicative of a full mystical experience by some authors (7). The version used in this study is under validation in French.

*6.1.1.3. Visual Analogue Scales (VAS)*

The VAS items included: “1: Any drug effect; 2: Good drug effect; 3: Bad drug effect; 4: I like the effect; 5: I am high; 6: I am stimulated; 7: I am scared; 8: Ego dissolution; 9: Happy; 10: Trust; 11: I feel close to others; 12: Talkative; 13: Open; 14: Speed of thought; 15: Perception of the speed of time; 16: Concentration.” For unidirectional items (1-8), participants marked their response along a scale ranging from “not at all” to “extremely”. Bidirectional items (9-16) included a “normal” midpoint, with anchors at either extreme.

Each VAS item was originally recorded as a score from 0 to 10. Unidirectional scales were rescaled to a 0-10 range, and bidirectional scales to a -10-10 range. For each participant, we calculated mean scores across sessions on each item and each principal component (PC), yielding individual PC1, PC2, PC3 values that could then be used in further analyses.

*6.1.1.4. First week dissociation scores*

At the SHU, dissociation was assessed by clinicians using the Clinician-Administered Dissociative States Scale (CADSS; scores from 0-92 points) (10), which was primarily administered during sessions 1 and 2. In contrast, at the CMME, dissociation intensity was rated by clinicians on a simpler 0-5 scale (0: no dissociation; 1: very slight; 2: slight; 3: moderate; 4: intense; 5: very intense). Therefore, to allow comparison across centres using different rating instruments, CADSS scores (0-92) were linearly rescaled to a 0-5 range. All available dissociation ratings from sessions 1 and 2 were included in the analyses.

*6.1.1.5. Calculation of total scores and summary measures*

For the MEQ, total scores were obtained by summing across all items and frequency of mystical experiences was calculated as the percentage of sessions in which MEQ-30 score ≥ 60. For the 5D-ASC and VAS, total scores were obtained by averaging across all items. For the CADSS, total scores were obtained by summing across all items. Whenever applicable, dimension scores were computed as the mean of their corresponding items. Per-participant mean scores across treatment were obtained by averaging across sessions. Peak intensity was defined as the highest score recorded for each participant.

**6.1.2. Power analysis**

A power analysis based on a prior report of correlations between MADRS change and mystical experiences (experience of unity: r = -0.440, p = 0.039, spirituality: r = -0.429, p = 0.039, insight: r = -0.398, p = 0.044) (11), indicated that n = 37 participants would provide 80% power to detect effects of comparable size. To account for effect size variability across trials and possible missing data, we decided to stop inclusion after 60 patients with a minimum of 8 esketamine sessions.

##### 6.1.3. Supplementary Statistical Analysis

##### 6.1.3.1. Outlier detection and missing data

Missing data were assessed for all variables prior to analysis. One participant was identified as a potential outlier due to a marked deterioration in depressive symptoms, showing a sudden increase of 25 points in MADRS scores between sessions 5 and 7, with missing data at session 6. A leave-one-out sensitivity analysis confirmed this observation (see Figure S3).

*6.1.3.2. Principal component analysis of subjective experience*

To identify data-driven dimensions of acute subjective effects induced by esketamine, principal component analyses (PCA) were applied separately to MEQ-30 items and VAS items, following approaches used in previous studies (12–16). PCA was used to characterise the structure of correlated (or anticorrelated) subjective effects without imposing predefined subscales. This approach was considered particularly relevant because the MEQ-30 was originally developed for classic psychedelics and may not optimally capture the phenomenology of esketamine-induced experiences.

Prior to PCA, each item was scaled to unit variance across participants, and items with zero variance were removed. Varimax rotation was applied to the component loadings (17).

The significance of principal components was assessed using permutation-based non-parametric testing. At each permutation, participant order was randomly shuffled for each symptom variable before recomputing the PCA. This procedure was repeated 10,000 times to generate a null distribution of explained variance. Components explaining a proportion of variance exceeding chance (p < 0.05) were retained for further analysis and ordered according to the variance explained.

A PCA including both MEQ-30 and VAS items combined was also conducted in the subset of participants who completed both scales (n = 29) (Figure S4). PCA was not applied to 5D-ASC scores because their collection was conditional on MEQ-30 score levels.

*6.1.3.3. Regression models and covariates*

Linear regression models were used to examine whether measures of subjective experience predicted antidepressant response (ΔMADRS = MADRS pre-treatment - MADRS post-treatment). All models controlled for age, sex, diagnosis (unipolar vs. bipolar), and treatment centre (SHU vs. CMME). Model assumptions of linearity, normality of residuals, and homoscedasticity were verified and met. Potential moderator effects of the covariates were tested via interaction terms; as none were significant, only main effects were retained in the final models and significance of predictors was assessed using Type II ANOVA.

To avoid multicollinearity and potential suppression effects arising from correlations between scale subdimensions, each dimension was tested in separate regression models and results for corrected for multiple comparisons.

Baseline WHOQOL spirituality scores and Big Five personality traits were also tested as predictors of both acute subjective experience scores (MEQ-30, VAS, dissociation) and antidepressant response in covariate-adjusted regression models.

Associations between changes in depressive symptoms measured with ΔQIDS and mystical-experience variables were examined using the same covariate-adjusted regression framework (see Supplementary Results).

*6.1.3.4. Pre-post treatment changes in WHOQOL spirituality and Big Five personality traits*

Changes in WHOQOL spirituality total scores from pre- to post-treatment were assessed using a paired t-test. Changes in Big Five personality traits were assessed using repeated-measures MANOVA, followed by corrected paired t-tests for significant effects. Pearson correlations were then used to test whether changes in personality correlated with antidepressant effects or mystical effects.

*6.1.3.5. Multiple comparison correction*

To control for multiple comparisons, family-wise error rates were adjusted using Bonferroni correction for small families of independent comparisons, including: (a) correlations between first-week mean dissociation scores and the four MEQ subdimensions, (b) correlations between first-week mean dissociation scores and the three VAS PCs, (c) regression models testing whether the three VAS PCs predicted ΔMADRS, (d) regression models testing whether baseline spirituality or personality measures predicted VAS PCs or MEQ dimensions, (e) comparisons of the four MEQ dimensions between mystical and non mystical groups, (d) t-tests testing changes in Big Five personality scores from pre- to post-treatment. For larger families of related tests, false discovery rates were controlled using the Benjamini-Hochberg (BH) procedure. This approach was applied to (a) comparisons of the 16 VAS item mean scores between mystical and non mystical groups, (b) correlations between the four MEQ dimensions and the three VAS PCs (15 tests).

*6.1.3.6. Bayesian statistics*

To quantify evidence in favour of the absence of effects, Bayesian analyses were conducted using the BayesFactor package. We report inverse Bayes factors (1/BF), representing the strength of evidence for the null hypothesis relative to the alternative. For example, 1/BF = 4 indicates that the data are four times more likely under the null hypothesis than under the alternative.

#### 6.2. Supplementary Results

*6.2.1. Supplementary Figures*

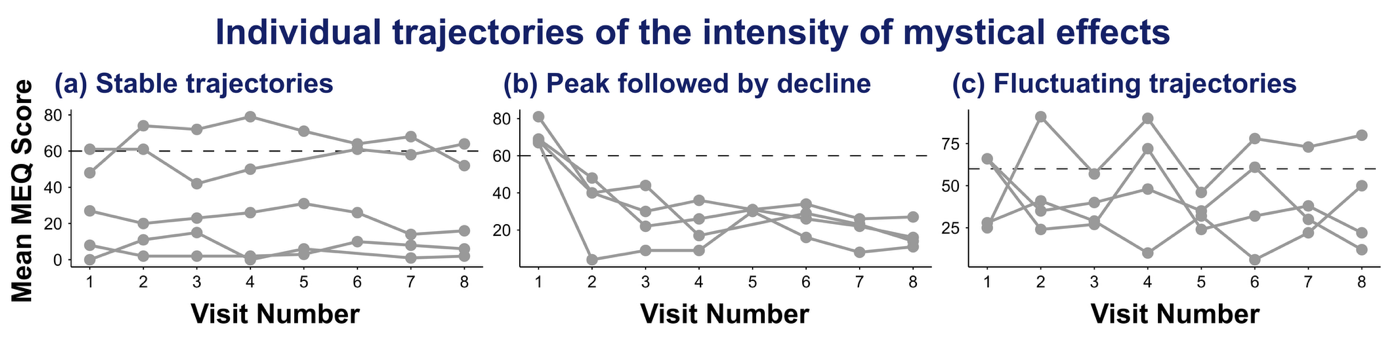
**Supplementary Figure S1.** Individual trajectories of MEQ scores across the 8 treatment sessions, grouped into three qualitative patterns: (a) stable trajectories, (b) peak followed by decline, and (c) fluctuating trajectories.

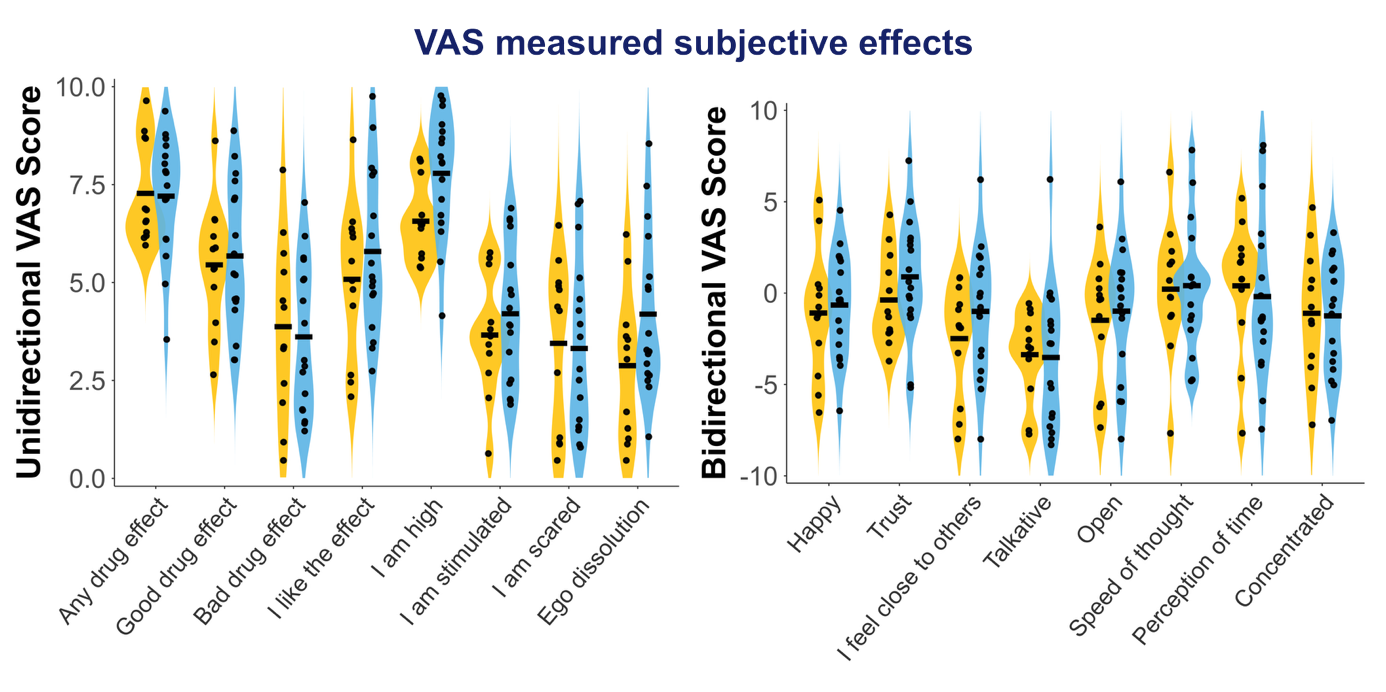
**Supplementary Figure S2.** Violin plots showing distributions of (left) unidirectional and (right) bidirectional VAS ratings in participants who completed the VAS scale (n = 29), split into mystical (blue) and non-mystical (yellow) groups. Group differences did not reach statistical significance after Benjamin-Hochberg (BH) false discovery rate (FDR) correction for multiple comparisons.

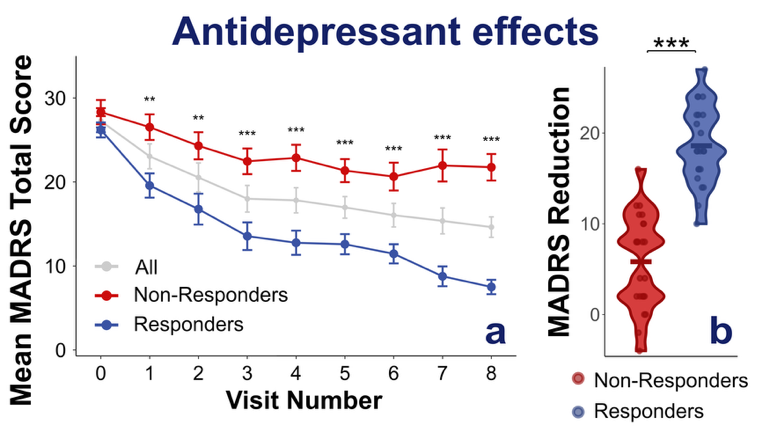

**Supplementary Figure S3**. (a) Trajectory of MADRS scores across sessions in the whole population (grey), non-responders (red), and responders (blue). Non-responders = <50% reduction in baseline MADRS score post-treatment; Responders = ≥50% reduction in baseline MADRS score post-treatment. (b) Violin plots showing the distribution of ΔMADRS (reduction in depression symptoms from pre- to post-treatment) in non-responders (red) and responders (blue).

*6.2.2. Participant flow*

57 patients were recruited, of which 12 were excluded: 3 discontinued treatment (1 due to nausea, 1 due to distressing anxiety, 1 at the end of hospitalisation); 1 was lost to follow up after being transferred to another unit; 2 withdrew consent while continuing treatment (1 due to anxiety from overanalysing experiences on the MEQ-30, 1 due to questionnaire fatigue); 2 switched to intravenous (IV) ketamine administration before finishing the induction phase; 3 had missing MADRS data for most sessions; and 1 was identified as an outlier as they showed a marked deterioration in MADRS scores between two sessions (sudden increase of 25 points between session 5 and 7 with missing data for session 6), likely due to external events.

*6.2.3. Esketamine treatment course*

Most patients received 8 biweekly sessions in the induction phase; 2 patients aged >65 received 9 sessions due to a lower first dose (28 mg vs 56 mg). For 3 patients, the final MADRS was taken after session 7 instead of 8, as assessment was not available after session 8.

In the CMME subgroup, 78% (18) started their first session at 84 mg, 17% (4) started at 56 mg and 1 (5%) started at 28 mg. In the SHU subgroup, 5% (1) started at 84 mg, 77% (17) started at 56 mg, and 18% (4) started at 28 mg. In total, 19 participants started at 84 mg, 21 started at 56 mg and 5 started at 28 mg. Esketamine dose in the first session did not predict ΔMADRS at the end of induction phase (r = -0.14, p = 0.37). Esketamine dose in the first session correlated with MEQ scores in the first session (r = 0.42, p = 0.004).

##### 6.2.4. Leave-one-out (LOO) analysis

One participant showed a marked deterioration in MADRS scores between two sessions (a sudden 25-point increase between sessions 5 and 7, with missing data for session 6), likely due to an external event. To confirm that this individual represented an outlier, we conducted a leave-one-out sensitivity analysis. In the full sample, mean MEQ scores were not a significant predictor of ΔMADRS after controlling for age, sex, diagnosis (unipolar vs. bipolar), and treatment centre (SHU vs. CMME) (*β* = 0.06, *p* = 0.29; Type II ANOVA). However, exclusion of the outlier markedly strengthened the association (*β* = 0.12, *p* = 0.029), whereas exclusion of any other participant had minimal impact (*|Δβ|* < 0.03, *p* > 0.2). Influence diagnostics (Cook’s distance = 0.50; DFBETA = −1.13) further confirmed that this participant exerted disproportionate influence on the relationship between MEQ scores and antidepressant response. Therefore, this participant was excluded from analyses (see Figure S1).

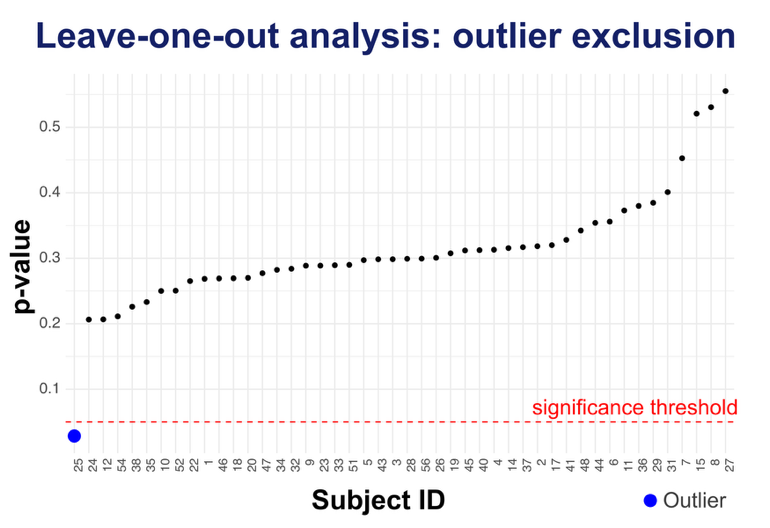

**Supplementary Figure S3.** Leave-one-out (LOO) analysis: p-values for the effect of mean MEQ scores on antidepressant response (regression model with covariates used in main analysis). Each point shows the p-value obtained when removing one participant and refitting the full model. Removing subject 25 (blue point) produced a significant effect (p < 0.05), indicating that this subject suppressed the association between MEQ and antidepressant response.

##### 6.2.5. Missing data

Eight patients had MADRS data missing in some sessions (in total: 10 of 362 sessions, ~2.8%). ΔMADRS (pre- to post-treatment) values were unaffected as the missing values did not occur at baseline or final sessions. Early ΔMADRS (from baseline to post session 2) had values missing for one subject.

Seven patients had MEQ data missing in some sessions (in total: 9 of 362 sessions, ~2.5%). MEQ mean, peak, and frequency were still calculated for these patients, as the missing data were minimal and likely missing at random.

2 (4.4%) patients had missing data for final WHOQOL, Big Five and QIDS assessments, as we could not contact them after the end of their treatment. Baseline QIDS was missing for one patient (2.2%).

Twenty-nine participants completed the VAS. Of those, 10 patients had VAS data missing in some sessions (in total: 9 of 234 sessions, ~3.8%).

Fifteen participants had missing first-week dissociation data and were excluded from the analysis (final n = 30).

##### 6.2.6. Mystical experiences predict self-reported antidepressant response (QIDS)

Across models, mystical experience measures significantly predicted ΔQIDS after controlling for age, sex, diagnosis, and centre. Higher mean MEQ scores (F(1, 36) = 6.47, p = 0.015) and higher peak MEQ scores (F(1, 36) = 7.56, p = 0.009) were each associated with greater symptom reduction, but frequency of mystical experiences was not (F(1,19) = 1.29, p = 0.27). Mean and peak MEQ scores in the first week also predicted reductions in QIDS scores from pre- to post-treatment (mean: F(1,36 = 10.04, p = 0.003; peak: F(1,36) = 8.16, p = 0.007).

*6.2.7. Mystical experiences still predict MADRS change when adjusting for baseline severity*

To account for potential regression-to-the-mean effects and baseline severity differences, we repeated the main analysis including baseline MADRS as a covariate (MADRS_final ~ PCs + MADRS_baseline + covariates), rather than modelling change scores. This adjustment did not substantially alter the results. Associations remained significant for mean MEQ scores (F(1,38) = 5.30, p = 0.027; previously F(1,39) = 5.17, p = 0.029) and peak MEQ scores (F(1,38) = 5.87, p = 0.020; previously F(1,39) = 5.91, p = 0.020), while MEQ frequency remained non-significant (F(1,20) = 3.09, p = 0.090; previously F(1,19) = 3.27, p = 0.090).

##### 6.2.8. Principal Component Analysis of MEQ-30 and VAS scores combined (n = 29)

We conducted a principal component analysis (PCA) on all MEQ-30 and VAS item scores across all sessions in the subset who completed both scales (n = 29). Four principal components (PCs) reached significance. PC1 primarily reflected the mystical dimension of the MEQ-30, accompanied by experiences of ecstasy and awe. PC2 captured the positive mood dimension of the MEQ-30, enjoyable drug effects measured by the VAS items “good drug effect”, “I like the effect”, “happy”, and social effects measured by the VAS items “trust”, “I feel close to others” and “open”. PC3 reflected the ineffability and transcendence of time and space MEQ-30 dimensions. PC4 represented a dysphoric, overstimulating experience. It loaded strongly on VAS items reflecting cognitive overstimulation - including increased speed of thought, perception of time, heightened concentration, and stimulation - as well as “ego dissolution” and “bad drug effect.” PC4 also showed moderate loadings on items related to social openness, such as “I feel close to others,” “talkative,” and “open” (Figure S4).

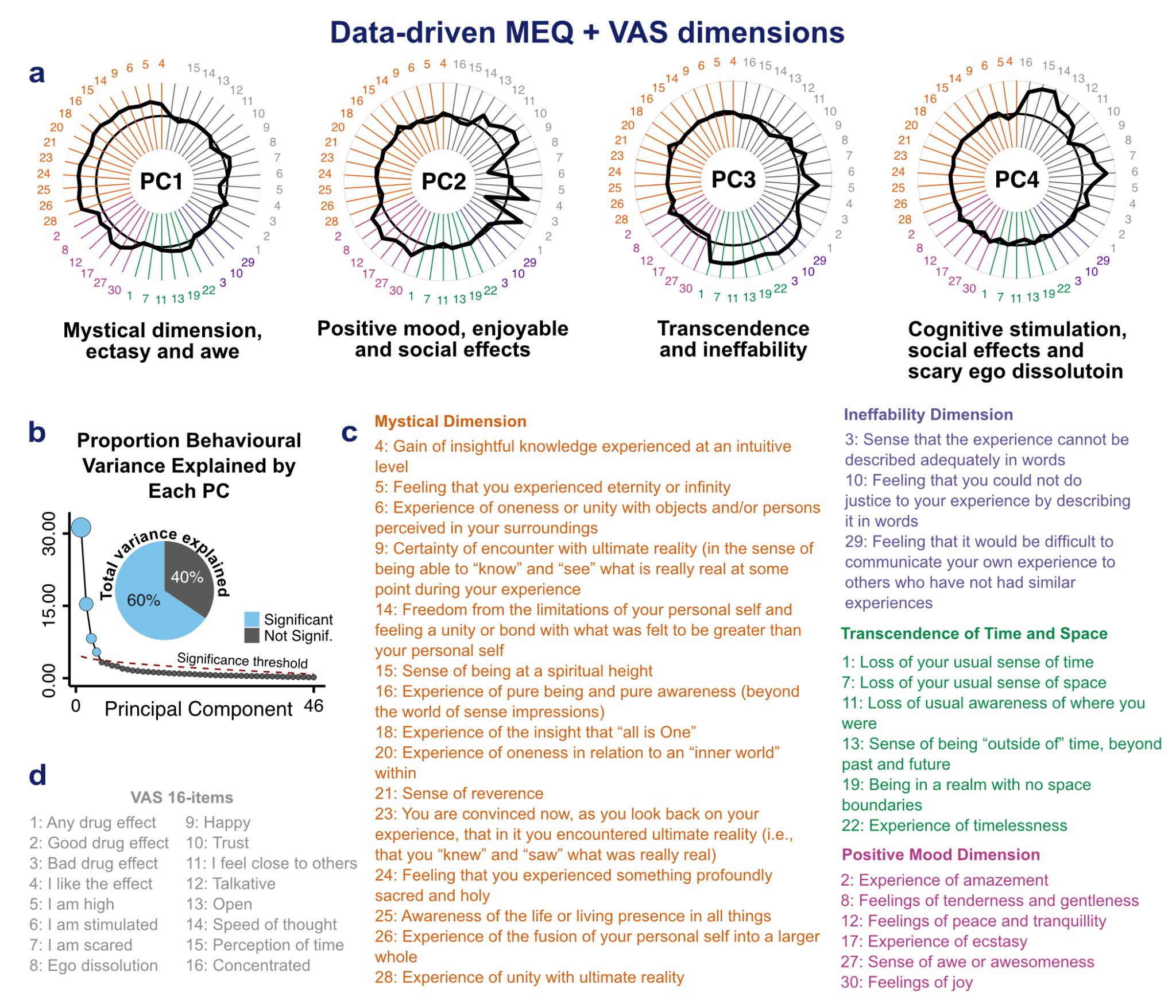

**Supplementary Figure S4.** Results of a principal component analysis (PCA) with MEQ-30 and VAS scores combined. PCA is a method that groups items that tend to co-vary to identify latent dimensions that explain variance in the data. (a) Radar plots showing the four principal components (PCs) and how strongly each MEQ-30 and VAS item contributes to them (loadings). These PCs represent the data-driven dimensions. (b) Scree plot showing how much of the total variance each PC explains. (c) MEQ-30 items grouped by their four established dimensions: Mystical, Positive Mood, Ineffability, and Transcendence of Time and Space. (d) The 16 VAS items included in the analysis.

*6.2.9. Pooled MEQ-30 + VAS PCs do not predict antidepressant effects*

None of the pooled MEQ-30 + VAS components significantly predicted changes in MADRS scores. Specifically, PC1 (F(1, 23) = 0.01, p = 0.92), PC2 (F(1, 23) = 1.61, p = 0.23), PC3 (F(1, 23) = 0.43, p = 0.52), and PC4 (F(1, 23) = 1.15, p = 0.29) were all non-significant in ANOVAs of linear regression models controlling for age, sex, diagnosis, and treatment centre (n = 29).

##### 6.2.10. Significant decreases in Neuroticism and Agreeableness post treatment

A repeated measures MANOVA revealed no significant main effect of time on the Big Five traits (Pillai's V = 0.022, F(1,42) = 0.90, *p* = 0.335). However, there was a significant main effect of trait (Pillai's V = 0.823, F(4,39) = 45.5, *p* < 0.001), and a significant time × trait interaction (Pillai's V = 0.402, F(4,39) = 6.60, *p* < 0.001). To explore which specific traits changed significantly from pre- to post-treatment, paired-sample t-tests were conducted for each trait. After correcting for multiple comparisons (Bonferroni correction with five comparisons), there was a significant decrease in Agreeableness (*ΔM* = -0.18, Bonferroni corrected *p* = 0.03), and in Neuroticism (*ΔM* = -0.25, Bonferroni corrected *p* = 0.02).

Interestingly, decreases in Neuroticism significantly correlated with MADRS decrease (*r* = 0.46, Bonferroni corrected *p* = 0.01) whereas decreases in Agreeableness did not (*r* = -0.25, Bonferroni corrected *p* = 0.5, 1/BF = 0.90), suggesting that clinical improvement is accompanied by a decrease of neuroticism only.

ΔNeuroticism and ΔAgreeableness did not correlate with MEQ scores (peak, mean or frequency), dissociation intensity, or VAS PC scores (all *ps* > 0.19, all 1/BF > 0.30), suggesting that personality changes occur independently of the nature of acute subjective effects.

##### 6.2.11. Baseline WHOQOL scores only predict peak 1^st^ week MEQ scores

Baseline WHOQOL scores did not significantly predict other MEQ scores, i.e. mean scores in the 1^st^ week, overall MEQ mean, overall peak or frequency of mystical experiences (all *ps* > 0.08 and 1/BF > 0.4).

**6.3. Supplementary Discussion**

#### *6.3.1. Esketamine antidepressant effects in relation with personality traits*

We observed a reduction in trait neuroticism from pre- to post-esketamine treatment, and this change correlated with antidepressant effects. This aligns with previous work showing that decreases in neuroticism often accompany decreases in depressive symptoms (18–21). Agreeableness also showed a small but significant decrease after treatment. Given that higher agreeableness is typically associated with lower depression (22,23), this pattern was unexpected. As this change did not correlate with antidepressant response, it may reflect a process occurring independently of depression improvement measured by MADRS. An alternative interpretation is that this result may reflect interpersonal changes not captured by MADRS, such as greater assertiveness.
